# Automated assessment of chest CT severity scores in patients suspected of COVID-19 infection

**DOI:** 10.1101/2022.12.28.22284027

**Authors:** Pranav Ajmera, Snehal Rathi, Udayan Dosi, Suvarna Lakshmi Kalli, Avinav Luthra, Sanjay Khaladkar, Richa Pant, Jitesh Seth, Pranshu Mishra, Manish Gawali, Yash Pargaonkar, Viraj Kulkarni, Amit Kharat

**Affiliations:** Dr. DY Patil Medical College, Hospital and Research Center, DY Patil Vidyapeeth, Pune, India; MGM Institute of Health Sciences, Navi Mumbai, India; DeepTek Medical Imaging Pvt. Ltd., Pune, India

## Abstract

**Background:** The COVID-19 pandemic has claimed numerous lives in the last three years. With new variants emerging every now and then, the world is still battling with the management of COVID-19.

**Purpose:** To utilize a deep learning model for the automatic detection of severity scores from chest CT scans of COVID-19 patients and compare its diagnostic performance with experienced human readers.

**Methods:** A deep learning model capable of identifying consolidations and ground-glass opacities from the chest CT images of COVID-19 patients was used to provide CT severity scores on a 25-point scale for definitive pathogen diagnosis. The model was tested on a dataset of 469 confirmed COVID-19 cases from a tertiary care hospital. The quantitative diagnostic performance of the model was compared with three experienced human readers.

**Results:** The test dataset consisted of 469 CT scans from 292 male (average age: 52.30 ± 15.90 years) and 177 female (average age: 53.47 ± 15.24) patients. The standalone model had an MAE of 3.192, which was lower than the average radiologists’ MAE of 3.471. The model achieved a precision of 0.69 [0.65, 0.74] and an F1 score of 0.67 [0.62, 0.71], which was significantly superior to the average reader precision of 0.68 [0.65, 0.71] and F1 score of 0.65 [0.63, 0.67]. The model demonstrated a sensitivity of 0.69 [95% CI: 0.65, 0.73] and specificity of 0.83 [95% CI: 0.81, 0.85], which was comparable to the performance of the three human readers, who had an average sensitivity of 0.71 [95% CI: 0.69, 0.73] and specificity of 0.84 [95% CI: 0.83, 0.85].

**Conclusion:** The AI model provided explainable results and performed at par with human readers in calculating CT severity scores from the chest CT scans of patients affected with COVID-19. The model had a lower MAE than that of the radiologists, indicating that the CTSS calculated by the AI was very close in absolute value to the CTSS determined by the reference standard.

## INTRODUCTION

COVID-19 has resulted in a substantial number of deaths in the past three years. The first case of COVID-19 was identified in December 2019 in Wuhan, China (1). The virus strain rapidly spread across the world, leading to the COVID-19 pandemic in March 2020 (2). While 81% of the people infected with COVID-19 develop minor to moderate symptoms such as mild pneumonia, 14% develop severe symptoms such as dyspnea, and 5% develop serious complications like respiratory failure, shock, or multiorgan dysfunction that can lead to death (3–5). Therefore, the sooner the patient is diagnosed, and supportive treatment is initiated, the lower the chances of disease progression (6).

Most COVID-19 management strategies involve symptomatic, emergency, and intensive care management as no specific drug against SARS CoV-2 has been developed yet (7). Frequent vaccination is the only method of prevention of COVID-19. Reverse Transcriptase Polymerase Chain Reaction (RT-PCR), a type of Nucleic Acid Amplification Test (NAAT), is the gold standard for the diagnosis of COVID-19 (8). According to the Fleischner society, diagnostic imaging such as chest radiography and Computed tomography (CT) plays a critical role in the diagnosis of COVID-19, particularly in moderate and severe categories and in individuals who have risk factors for disease progression (9). While the choice between imaging modalities is at the discretion of the physician, CT is particularly effective in establishing a baseline of the disease. The morphological pattern in the lungs observed on a CT scan is associated with the progression and severity of COVID-19 infection. Ground-glass opacities (GGOs), air bronchogram, consolidation, crazy paving pattern, and interlobular septal thickening with bilateral and multi-lobe involvement are the most common radiological features associated with COVID-19 observed in chest CT scans (10–13). The CT-Severity Score (CTSS) is used to determine the severity of lung involvement depending on the percentage of lung parenchyma affected. Various scoring systems have been proposed and implemented in the past. Li et al. used a 25-point scoring system based on the percentage involvement of each lobe of the lung (14). Although 96-point and 40-point scoring systems are available to score the severity of lung involvement in COVID-19 patients, the 25-point scoring system is the most widely used, and it was first devised and popularized by Chang et al. in 2005 for assessing the severity of Severe Acute Respiratory Syndrome (SARS) (15). Depending on the severity score and clinical and biochemical parameters, decisions concerning home-based care, hospital ward-based treatment, or Intensive Care Unit (ICU)-based treatment can be made (16,17). However, the high caseload causes a diagnostic backlog, increases the likelihood of medical error, and contributes significantly to burnout among clinicians (18).

Artificial Intelligence (AI) can play an important role in aiding the diagnosis of the severity of COVID-19. AI models can help the radiologist in assessing CT scans and providing information on the extent of lung involvement. The AI model that quantifies the severity of lung involvement is of great assistance to clinicians in the field of pulmonology and emergency medicine to begin therapy immediately. Herein, we validate the performance of an AI model for automatic prediction of the COVID-19 severity score. The model can identify the scan as positive for COVID-19 and subsequently categorize it based on severity through the quantification of CTSS. We evaluate the model’s performance and compare it with the performance of board-certified readers.

## MATERIALS AND METHODS

### Dataset

This was a retrospective study approved by the Institutional Ethical Committee (IEC) of a tertiary care hospital and research center. All patients provided informed consent to undergo the CT scan procedure. The patient data was acquired in accordance with the Health Information Portability and Accountability Act (HIPAA) to ensure patient privacy and anonymity. All the patients who underwent a CT thorax examination during the study period to either rule out COVID-19 or reassess the status of COVID-19 were included in the study. Strict inclusion and exclusion criteria were implemented to include scans that showed features suggestive of COVID-19 infection. Abnormalities that were considered significant for the diagnosis of disease included the following: ground-glass opacities, consolidation, crazy paving pattern, nodules, reticulation, and subpleural curvilinear line in a predominantly peripheral distribution (19). Patients who had a positive RT-PCR test prior to 14 days of the CT examination or had a positive RT-PCR test result within 3 days after the CT scan procedure, were included in the study. Following the inclusion and exclusion criteria, the final dataset included 469 chest CT scans. Figure 1 illustrates an overview of participant selection and patient inclusion and exclusion criteria in the external test set.

**Figure 1:**
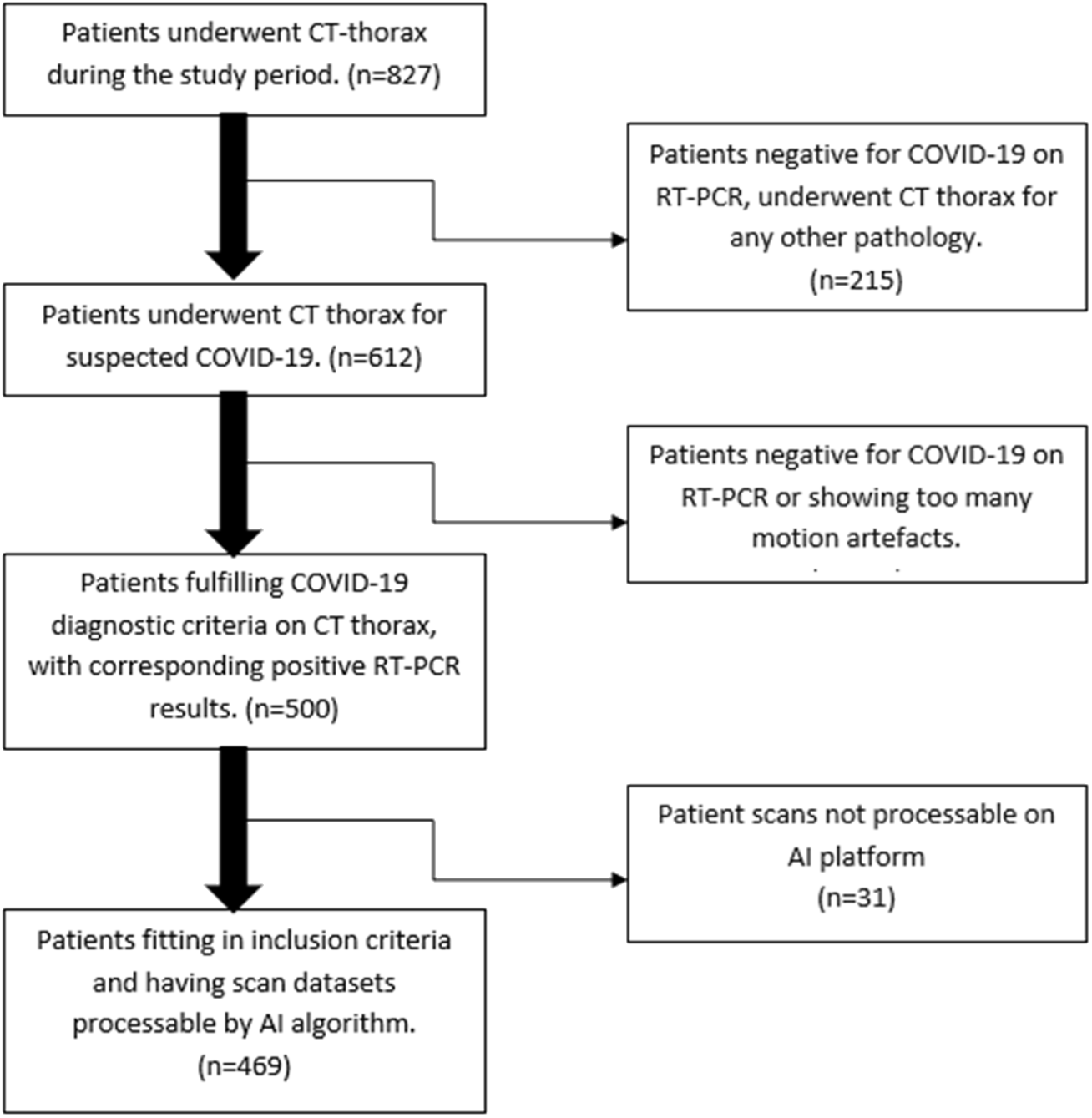
Flowchart illustrating the external validation CT datasets used in the study after implementing inclusion and exclusion criteria.

### Image acquisition

The CT thorax examination was performed in all patients using a Siemens Somatom-CT 16 slice scanner (Siemens Medical Solutions, USA) at full inspiration from the thoracic inlet to the costophrenic angle level with the following parameters: tube voltage of 120kVP, tube current of 110mAs, the pitch of 1.3mm, and the scan time of 23.5s.

### Establishment of Ground Truth

To establish the ground truth, all COVID-19 CT scans were annotated by two radiologists with a combined experience of 32+ years. Lung lobe-wise and the total COVID-19 severity score for each patient was calculated by the annotating radiologists. For this study, we followed the 25-point severity score classification system. Each of the five lung lobes was scored on a scale of 0 to 5 wherein, 0: no lobe involvement, 1: < 5% lobe involvement, 2: 5% - 25% lobe involvement, 3: 26% - 49% lobe involvement, 4: 50% - 75% lobe involvement, and 5: >75% of lobe involvement. The total CT severity score is the sum of the individual lobe scores and ranges from 0 (no lung involvement) to 25 (maximum lung involvement). Based on the severity score, each case was categorized as mild (CTSS: 1-7), moderate (CTSS: 8-17), or severe (CTSS >18).

### AI model

All scans were de-identified and processed using DeepTek’s cloud-based AI platform. The model used Convolutional Neural Networks (CNNs) to identify consolidation and GGOs on each slice of the scan. The CNN parameters were optimized using a proprietary algorithm. Each CT slice was resized and normalized to standardize the acquisition process. It was then used as an input to the AI model, which quantifies the extent of infection in each lobe. It then converts this quantity to a lobe-wise severity score (an integer between 0-5). The severity scores of all lobes are added to give the final output — a total CT severity score between 0 and 25.

### Comparison of the diagnostic performance of the deep learning model and human readers

The chest CT scans from 469 patients with positive RT-PCR tests were used for comparing the diagnostic performance of the model with three experienced readers (R1, R2, and R3). R1, R2, and R3 participated as test readers and were not involved in the establishment of ground truth for this study. R1 was an emergency medicine clinician who had been working as the first responder in all cases reporting to the casualty throughout the period of the COVID-19 pandemic and had an experience of 5 years. R2 was a consultant radiologist with an experience of 4 years who had been reporting COVID-19 CT scans regularly throughout the pandemic. R3 was a consultant pulmonologist who worked in the respiratory ICU during the COVID-19 pandemic and had an experience of 4 years.

The participating readers were informed that all the scans were from the patients who tested positive for COVID-19 on RT-PCR and were instructed to assess the COVID-19 CTSS of individual lobes and scan using the 25-point severity scoring system.

### Statistical Analysis

All analysis and computation were performed in python using SciPy statistical library (version 1.4.1). The comprehensive evaluation of the model and radiologists’ performance on the test set included MAE, MSE, sensitivity, specificity, precision, and F1 score. While the MAE and MSE were calculated on the total CTSS, all other metrics were calculated using the macro-average of the metrics in the three classes: mild, moderate, and severe. To measure the variability in these values, we used 95% confidence intervals using the empirical bootstrapping method. To compare the difference in the average performance of readers and the AI model, Wilcoxon signed-rank test was performed. The alternative hypothesis for MAE and MSE significance testing assumed that the AI metric had a lower value than the average human reader metric. The alternative hypothesis for significance testing of all other metrics assumed that the performance of AI is superior to the performance of the average human reader. A p-value of less than 0.05 was considered statistically significant. To assess the agreement in CTSS reporting between the AI and readers, Bland-Altman (B-A) analysis was performed.

## RESULTS

### Patient population and clinical features

Table 1 summarizes the study population characteristics for the independent clinical test set. The test dataset included 469 scans from 292 male patients (mean age: 52.30 ± 15.90 years) and 177 female patients (mean age: 53.47 ± 15.24 years). The age distribution of patients enrolled in this study is represented in Supplementary Fig 1. The most prevalent symptoms in the study population at the time of the CT examination were cough and fever. 18.9% of patients had at least one of the following comorbidities: hypertension (HTN), diabetes mellitus (DM), tuberculosis (TB), and bronchial asthma (BA).

**Table 1:** Demographics and characteristics of patients with COVID-19

| Characteristic | Number of patients (%) |
| --- | --- |
| **Patient information** |  |
| Total no. of participants | 469 |
| No. of female participants | 177 (37.7) |
| No. of male participants | 292 (62.3) |
| **Signs and symptoms** |  |
| Breathlessness | 159 (33.9) |
| Cough | 270 (57.5) |
| Fever | 273 (58.2) |
| Body ache | 24 (5.1) |
| **Comorbidities** |  |
| DM, HTN, TB, BA | 89 (33.1) |

### Performance of deep learning model in assessing CT severity scores

The CTSS is categorized as mild, moderate and severe, based on the 25-point visual grading system proposed by Li et al. (14). They recommended assigning a score of 0-5 to each lobe, based on the extent of involvement of each lobe: 0: 0% involvement; score 1: less than 5% involvement; score 2: 5% to 25% involvement; score 3: 26% to 49% involvement; score 4: 50% to 75% involvement; score 5: >75% involvement. Therefore, each lobe can have a score between 0 to 5.

The total score is calculated by adding the scores of each lobe, and the CT scan is classified into either of three categories: scans with a total score of <=7 as mild, 8-17 as moderate, and >=18 as severe (20). Table 2 represents the grading discrepancies between the AI model and the human readers. The model showed superiority in grading mild cases of COVID-19 infection. For grading moderate cases of COVID-19, the model performed equivalent to the readers.

**Table 2:**
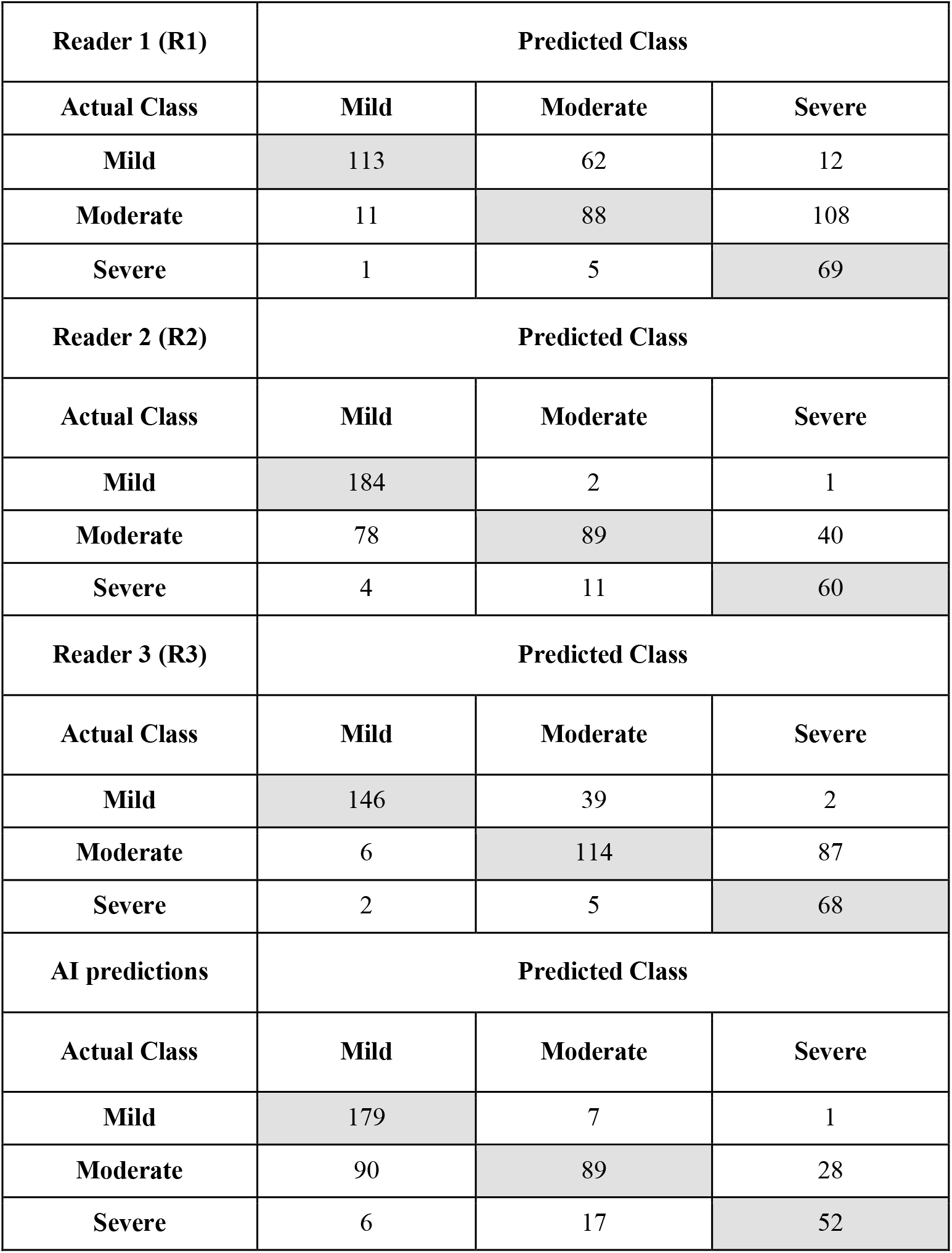
Confusion matrix comparing the grading performance of CT severity scores between AI model and experienced human readers.

### Model output

The output of the model included masks for consolidation and GGOs. The model provided an explainable approach for the identification of the severity of infection. Fig 2 shows the representative CT images of a COVID-19-affected patient and the corresponding outputs of the deep learning model.

**Figure 2:**
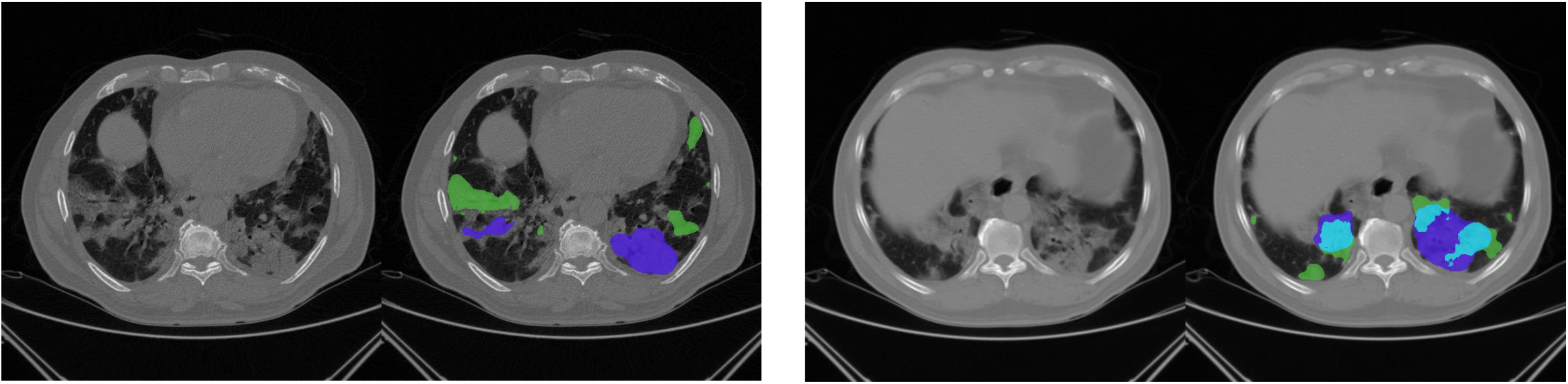
Slices (original and with AI-predicted masks) from a CT scan of a 65-year-old male patient affected with COVID-19. The AI masks highlight the regions with consolidation (blue) and GGO (green). Light blue indicates a prediction of both consolidation and GGO within the same region.

### Comparison of performance between deep learning model and experienced radiologists

The performances of the deep learning model and human readers in assessing severity scores from chest CT scans are presented in Table 3. The model had a mean absolute error (MAE) of 3.192 [2.965, 3.435] and a mean squared error (MSE) of 16.953 [14.18, 20.32], which were significantly lower (p=0.002) than the average radiologists’ MAE of 3.471 [3.324, 3.617] and MSE of 20.031 [18.17, 22.09]. A lower MAE indicates that the CTSS calculated by the AI model were very close in absolute value to the CTSS determined by the reference standard. The model achieved a precision of 0.69 [0.65, 0.74] and an F1 score of 0.67 [0.62, 0.71], which was significantly superior to the average reader precision of 0.68 [0.65, 0.71] and average reader F1 score of 0.65 [0.63, 0.67]. The model demonstrated a sensitivity of 0.69 [0.65, 0.73] and specificity of 0.83 [0.81, 0.85], which was comparable to human readers’ sensitivity of 0.71 [0.69, 0.73] and specificity of 0.84 [0.83, 0.85].

**Table 3:** Performance of the deep learning model vs human readers

| Metrics | R1 | R2 | R3 | Average of R1, R2, and R3 | Standalone AI | p-value |
| --- | --- | --- | --- | --- | --- | --- |
| **MAE**<br>[95% CI] | 4.18 [3.90, 4.46] | 3.33 [3.09, 3.57] | 2.90 [2.67, 3.13] | 3.47 [3.32, 3.62] | 3.19 [2.97, 3.44] | 0.002 |
| **MSE**<br>[95% CI] | 27.26 [23.58, 31.45] | 17.76 [14.88, 21.17] | 15.08 [12.35, 18.33] | 20.03 [18.17, 22.09] | 16.95 [14.18, 20.32] | 0.002 |
| **Sensitivity**<br>[95% CI] | 0.65 [0.61, 0.69] | 0.74 [0.69, 0.78] | 0.75 [0.71, 0.78] | 0.71 [0.69, 0.73] | 0.69 [0.65, 0.73] | 0.059 |
| **Precision**<br>[95% CI] | 0.61 [0.57, 0.65] | 0.72 [0.68, 0.76] | 0.70 [0.66, 0.74] | 0.68 [0.65, 0.71] | 0.69 [0.65, 0.74] | 0.005 |
| **Specificity**<br>[95% CI] | 0.79 [0.78, 0.82] | 0.85 [0.83, 0.87] | 0.86 [0.84, 0.88] | 0.84 [0.83, 0.85] | 0.83 [0.81, 0.85] | 0.386 |
| **F1 Score**<br>[95% CI] | 0.58 [0.53, 0.62] | 0.69 [0.65, 0.73] | 0.69 [0.65, 0.73] | 0.65 [0.63, 0.67] | 0.67 [0.62, 0.71] | 0.007 |

The Bland-Altman (B-A) analysis was used to demonstrate agreement between the readers and the AI model in calculating CTSS. A mean difference value close to zero represents that the AI model performed well in comparison to the average of human readers. Our analysis demonstrated a mean agreement difference of -3.154 (95% CI: -9.171, 2.863) for the test dataset (Fig. 3). The B-A analysis for the AI model and individual readers is presented in Supplementary Fig. 2.

**Figure 3:**
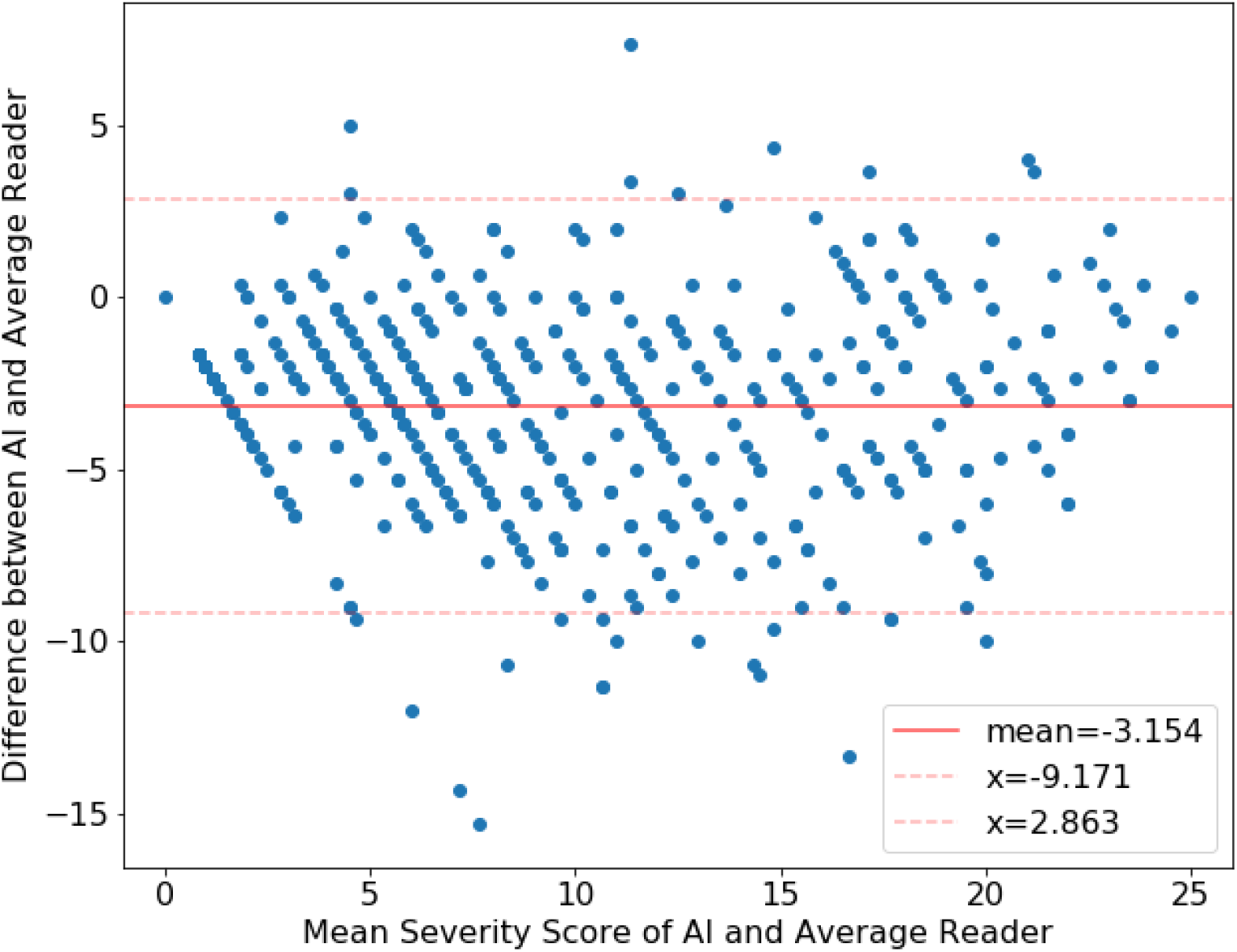
B–A plot illustrating the agreement test for CTSS calculation between the readers and the AI model. The plots elucidate the difference in the CTSS reported by the readers and calculated by the AI model (y-axis) over the mean CTSS reported by the readers and calculated by the AI model (x-axis). The value of the mean difference (red solid line) and the upper and lower limits of 95% agreement (red dotted lines) are shown.

## DISCUSSION

The American College of Radiology (ACR) recommends the use of computed tomography for suspected COVID-19 infection (21). However, visual assessment of COVID-19 lesions from chest CT scans is time-consuming, suffers from inter- and intra-reader variability, and lacks reproducibility (22). Due to these factors and the high workload experienced by medical practitioners, a semi-quantitative visual assessment of COVID-19 is impractical in clinical practice. Computer-aided diagnosis (CADx) has emerged as an important adjunct tool in diagnostic imaging. As artificial intelligence has advanced, deep learning models have started to be frequently employed for the detection and diagnosis of clinical conditions.

Several research groups have been working on COVID-19 detection and diagnosis using AI tools on radiological exams. Ghaderzadeh & Asadi (23) reviewed numerous studies and identified that classification models are most commonly used for COVID-19 detection on CT scans and X-rays. While COVID-19 can be diagnosed using different radiological exams, including chest radiography (24)(25) and lung ultrasounds (26), computed tomography (CT) have the highest sensitivity (8). Moreover, CT severity scoring goes beyond detection; it is an accurate measure of severity that aids in proper disease prognosis and treatment. CT scans aid in lobar analysis, which provides detailed information about COVID-19 disease progression and associated mortality risk (27). Some studies attempted to calculate severity scores, but there is no universal scoring system or performance metric for scoring the severity of COVID-19. While Kardos et. al. (28) also used a 0-25 prediction system, the only inference they drew was the presence or absence of COVID-19. Shiri et al. (29) used a severity scoring system with only four classes. These classes did not consider the individual lobe involvement in the disease but included other clinical symptoms. Neither of these two studies reported the MAE for their algorithms. In a deep learning task with numerical outcomes, MAE provides a notion of how much the results will vary in clinical settings. Users can compare it with the range of severity levels (e.g. 0-25) and decide whether it is acceptable.

While previous studies (28–31) calculated metrics using a range of severity scores from 0-3 to 0- 6, we employed a 25-point severity scoring system. We did an extensive survey of a deep learning model on a hospital dataset with performance that matched three experienced human readers. The model used in this study can identify features suggestive of COVID-19 and calculate COVID-19 CT severity scores based on that. By highlighting consolidations and GGOs within the CT slice, the model provided an explainable solution for assigning a severity score to the CT scan. This would provide radiologists with insight into the model’s decision-making process. The specificity and sensitivity of the model were comparable to the average performance of the panel of readers from various backgrounds, whereas the precision and F1 score of the model were significantly better than the readers. The model exhibited an MAE of 3.192 which was lower than the mean MAE of 3.471 exhibited by the expert readers, indicating that the CTSS predicted by the model were very close in absolute value to the CTSS determined by the reference standard. The automated CTSS calculation was in good agreement with CTSS assessed by experienced readers, as demonstrated by Bland-Altman analysis. The AI model used in the study was comparable in performance to three expert board-certified human readers and can be utilized in clinical practice.

This retrospective study had certain limitations. First, the study was single-centric, which partly limits its generalizability to larger populations. Second, while the ground truth consisted of the consensus opinion of two radiologists, it would have been ideal to have more radiologists to further minimize reader bias.

In conclusion, the model used in the study helped in categorizing patients with COVID-19 based on the severity of the disease. The model performed equivalent to human readers in calculating CTSS from the chest CT scans of patients affected with COVID-19 and demonstrated a low MAE indicating that the CTSS calculated by the AI model was close to the CTSS determined by the reference standard. In clinical practice, this model could reduce the turnaround time for evaluating COVID-19 cases and help in triaging patients based on the severity scores. This would help reduce the burden on an already overburdened healthcare system.

## Data Availability

All data produced in the present study are available upon reasonable request to the authors

