## Supplementary Figures for "Automated assessment of chest CT severity scores in patients suspected of COVID-19 infection"

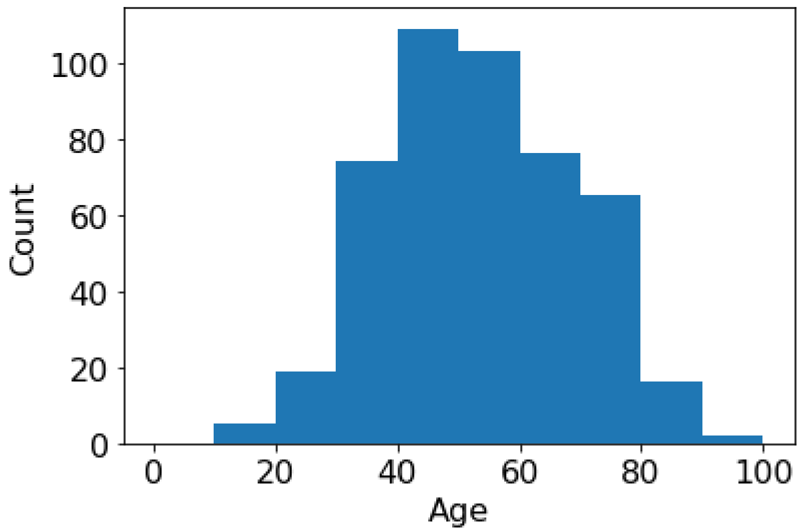


**Supplementary Figure 1:** Age distribution of COVID-19-affected patients.

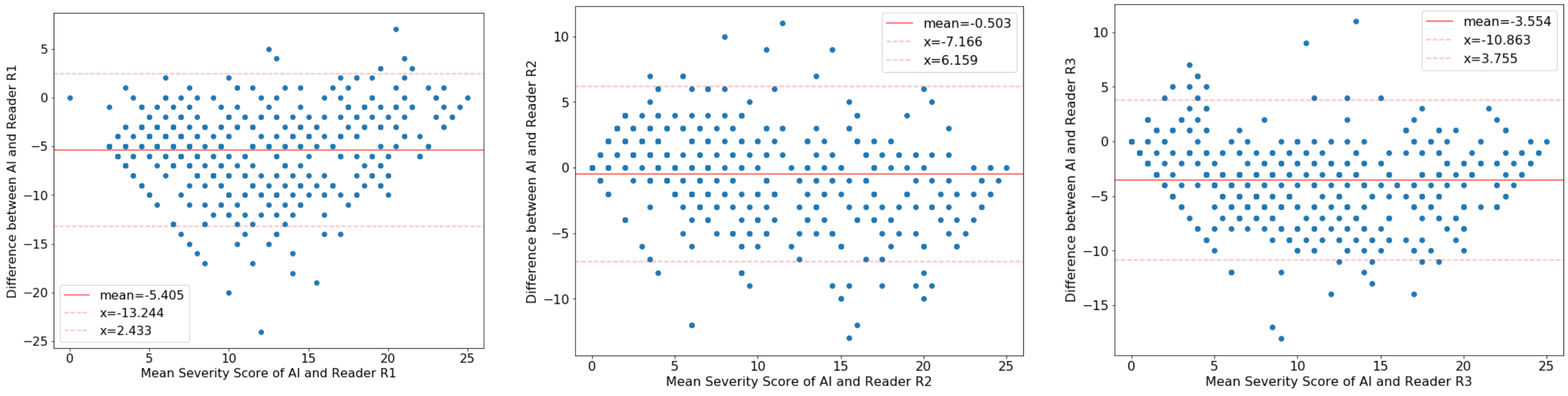


**Supplementary Figure 2:** B–A plot illustrating the agreement test for CTSS calculation between the individual readers and the AI model. The plots elucidate the difference in the CTSS reported by the individual reader and calculated by the AI model (y-axis) over the mean CTSS reported by the individual reader and calculated by the AI model (x-axis). The value of the mean difference (red solid line) and the upper and lower limits of 95% agreement (red dotted lines) are shown.
